# Locus-restricted genetic overlap between pelvic organ prolapse and estimated bone mineral density at 18q21.1 (SMAD7), with unresolved regulatory direction

**DOI:** 10.64898/2026.11.24.26368921

**Authors:** Jinhua Tian, Hongming Li, Songming He

## Abstract

Pelvic organ prolapse (POP) is highly prevalent in parous women. Observational studies link POP with low bone mineral density (BMD), but shared genetic determinants are unclear. Global Mendelian randomization has not supported a causal effect of BMD on POP. We therefore asked whether overlap exists at specific loci. We merged 2,524 genome-wide significant FinnGen R12 POP variants (23,074 cases, 130,160 controls) into 30 loci, seven of which lay within 500 kb of an instrument for estimated BMD (eBMD). The clearest example is 18q21.1: lead variant rs4939832 (G allele) was associated with POP (β = 0.071, P = 9.5 × 10^−9^) and with eBMD (β = 0.018 per SD, P = 1.3 × 10^−11^) at the same allele. Regional colocalisation gave a posterior probability of 0.98 for signal sharing under the single-causal-variant assumption. LD-aware fine-mapping then resolved two adjacent independent causal signals with no overlapping credible sets (rs4939832 versus rs2337106; r^2^ = 0.21). We then queried rs4939832 against SMAD7 in 28 blood and immune-cell eQTL analyses. No analysis reached nominal significance (minimum P = 0.118; pooled P = 0.78). Analyses within a panel are not statistically independent. Interrogating the closest target-adjacent tissues (GTEx vagina, n = 141; uterus, n = 129) likewise returned no significant cis-association (P = 0.26 and 0.77), so any cis-regulatory effect of this variant is likely small and its directionality cannot be reliably inferred from available tissue resources. Supporting evidence from independent cohorts was limited: the variant showed only nominal association in UK Biobank POP (P = 0.058) and was not assayed in the largest POP GWAS. POP and eBMD therefore overlap in a locus-restricted manner rather than through a systemic bone-mass pathway, and the cis-regulatory direction of rs4939832 remains unresolved. Functional interpretation requires well-powered eQTL data from genuine pelvic-floor connective-tissue compartments, which are not publicly available.

**Author Summary:** Pelvic organ prolapse is a common condition in which pelvic organs shift downward, causing discomfort and often requiring surgery. Women with prolapse tend to have lower bone mineral density, but whether the two conditions share a common genetic cause has been unclear. Large genetic studies had already ruled out a genome-wide causal effect of bone density on prolapse. We therefore focused on individual genomic regions and found one region, 18q21.1, where the same genetic variant (rs4939832) is associated with both higher prolapse risk and higher estimated bone density. Advanced statistical fine-mapping methods resolved two distinct neighbouring genetic signals instead of a single shared causal variant. We then asked whether the prolapse variant changes the activity of the nearby SMAD7 gene, testing 28 blood-based datasets and, as the closest available tissues, vagina and uterus from the public GTEx resource. No detectable effect was found in any of them. The overlap between the two traits therefore appears confined to this single chromosome region rather than reflecting a body-wide bone pathway, and functional interpretation must wait for studies of genuine pelvic-floor tissue.

## 1. Introduction

Pelvic organ prolapse is highly prevalent. Symptom-based prevalence is 3–6%, and up to 50% of parous women show prolapse on vaginal examination [1]. Connective-tissue integrity is central to pelvic floor support. Moderate-to-severe prolapse, particularly rectocele, was associated with fracture history and lower whole-body BMD in the Women’s Health Initiative trial [2]. These observations motivated a search for shared genetic determinants, although the observational association is subtype-specific and may partly reflect shared age structure.

Two complementary genetic designs can address shared determinants. Global Mendelian randomization (MR) tests whether genetically predicted BMD causally affects POP across the genome. Locus-level comparison asks whether specific regions carry signals for both traits. A companion two-sample MR analysis using 517 independent eBMD-derived instruments (see S5 Table) found no causal effect of eBMD on POP (OR = 1.01 per SD, P = 0.73), with a formal equivalence bound of ±10% per SD. The global answer is therefore negative.

The locus-level question remained open. GWAS have identified risk loci for POP [3], and a large GWAS atlas has mapped hundreds of loci for estimated BMD [4]. Comparing the two maps at matched loci is a direct test of locus-restricted overlap. Here we report the clearest example that emerged from this comparison: chromosome 18q21.1, where the POP lead variant rs4939832 lies within SMAD7, a well-replicated eBMD locus encoding an intracellular inhibitor of TGF-β signalling [4,5].

In this work, we evaluated whether 18q21.1 harbours aligned association signals for POP and eBMD, and whether observed overlap arises from one shared causal variant or two neighbouring independent variants. We further investigated whether rs4939832 modulates SMAD7 expression, as the simplest biological model would predict. For the regulatory question we used all publicly available blood and immune-cell eQTL resources and report their null result with explicit attention to the non-independence of contributing datasets.

## 2. Results

### 2.1 A single genome-wide significant variant at 18q21.1 in FinnGen

The FinnGen R12 endpoint N14_FEMGENPROL (female genital prolapse) comprised 23,074 cases and 130,160 controls (case–control effective sample size 78,398) [6]. In the released summary statistics we counted 2,524 variants at P < 5 × 10^−8^, which merged within ±500 kb into 30 loci. Seven POP loci lay within 500 kb of an eBMD instrument. The 18q21.1 locus was represented by a single genome-wide significant variant, rs4939832 (GRCh38 chr18:48,939,295; A > G; effect allele G; G-allele frequency 0.255). Its association with POP was β = 0.071 per G allele (SE 0.0124; z = 5.74; P = 9.5 × 10^−9^; Table 1).

**Table 1.** Association of rs4939832 (G allele) with POP and estimated BMD.

| Dataset | Phenotype | n | Cases | Controls | β (per G) | SE | z | P | EAF (G) |
|---|---|---|---|---|---|---|---|---|---|
| FinnGen R12 (GRCh38) | Female genital prolapse (N14_FEMGENPROL) | 153,234 | 23,074 | 130,160 | +0.0712 | 0.0124 | 5.74 | 9.5 × 10<sup>−9</sup> | 0.255 |
| Morris et al. 2019 (GRCh37) | eBMD, heel quantitative ultrasound (SD units) | 426,824 | — | — | +0.0176 | 0.0022 | 8.07<sup>a</sup> | 1.3 × 10<sup>−11</sup> | 0.242 |
| UKB round 2, Neale lab (GRCh37) | Genital prolapse (ICD-10 N81; both sexes; linear scale) | 361,194 | — | — | +7.43 × 10<sup>−4</sup> | 3.91 × 10<sup>−4</sup> | 1.90 | 0.058 | 0.2426 |

β values are per effect allele (G) and are not directly comparable across studies because the phenotypes and models differ; only z-scores and directions are compared. Case and control counts for the UKB and eBMD resources follow the respective publications. For FinnGen, n = 23,074 + 130,160 = 153,234. <sup>a</sup> z values are computed as β/SE and are used only to place the three estimates on a common scale. For the eBMD row, the P value is the one reported by Morris et al. In the released file this reported P is not exactly reproducible from β and SE for strong signals, so published P values were retained for inference throughout.

### 2.2 The same allele is associated with higher estimated BMD

We extracted the corresponding statistics from the eBMD GWAS of Morris et al. (UK Biobank heel quantitative ultrasound; n = 426,824) [4] after harmonising alleles to the FinnGen ALT strand. The G allele of rs4939832 was associated with higher eBMD: β = +0.0176 per SD (SE 0.0022; P = 1.3 × 10^−11^ as reported; G-allele frequency 0.242; Table 1). The direction of effect was therefore the same for both traits. Fig 1 shows the z-score profiles of the two traits across the 1.6 Mb region. The eBMD signal in the window is broader than the POP signal. Its maximum |z| (9.94) is attained by a variant other than rs4939832, so the two lead signals are close but not identical. This motivated follow-up colocalisation and fine-mapping to disentangle signal relationships within the locus.

**Figure 1.**
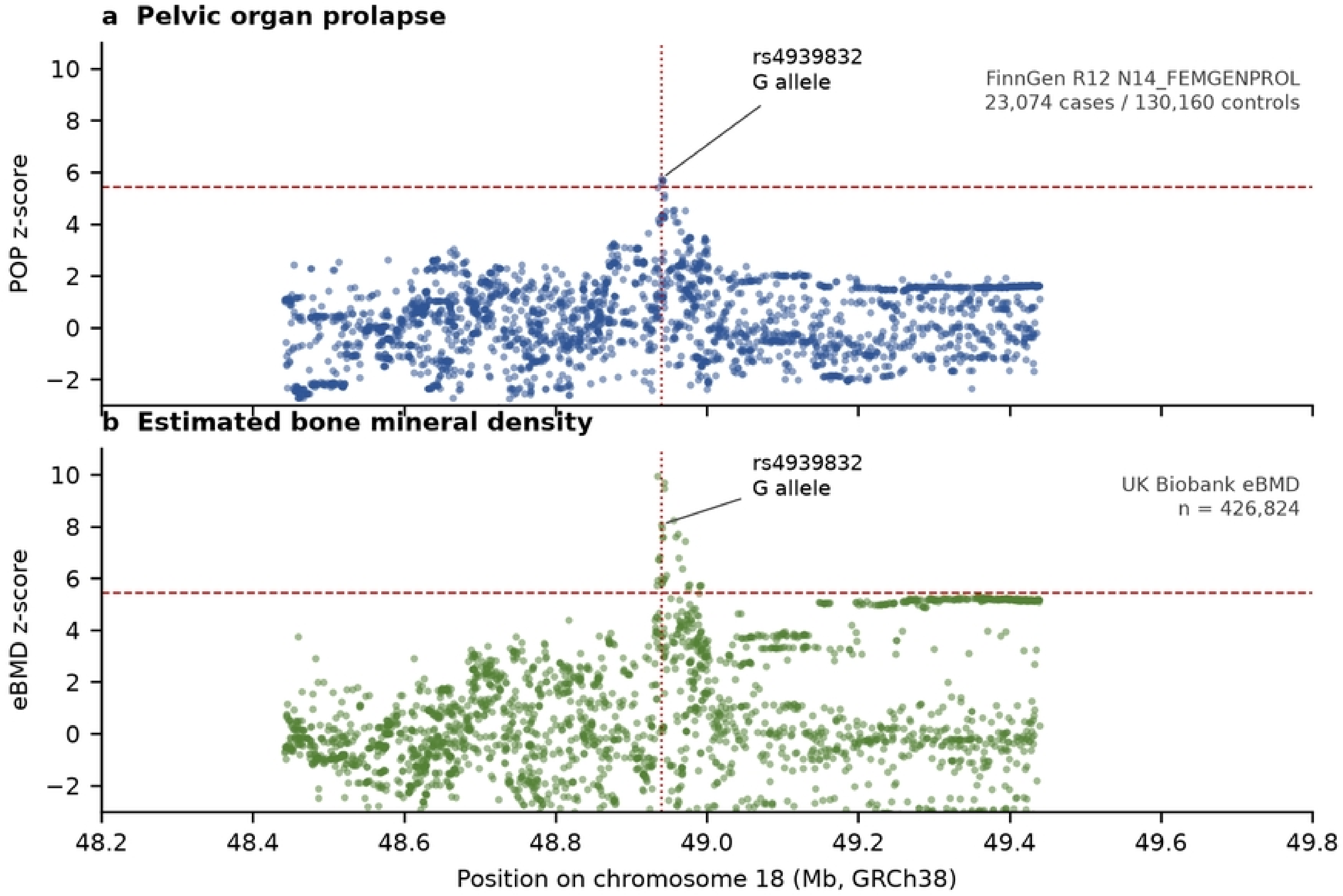
Aligned z-score profiles at 18q21.1 for POP and eBMD. (a) FinnGen R12 female genital prolapse. (b) Estimated BMD from Morris et al. (UK Biobank). Points are variants in the 1.6 Mb region (GRCh38 chr18:48.2–49.8 Mb) present in both datasets. z-scores are computed as β/SE and are used here only to display signal location and relative strength. Dashed lines mark the nominal genome-wide significance level (z ≈ 5.45). The dotted vertical line marks rs4939832. The eBMD window maximum (|z| = 9.94) is attained by a variant other than rs4939832.

### 2.3 Regional colocalisation, resolved by fine-mapping into two adjacent signals

The coloc.abf method assumes exactly one causal variant and does not model linkage disequilibrium. Regional colocalisation with coloc.abf using 2,491 variants across the locus gave a posterior probability of 0.98 for hypothesis H4 (one shared causal variant) and 0.02 for H3 (two distinct linked variants) [7]. Because two nearby signals joined by weak LD can inflate H4 under this assumption [7], we re-examined the locus with LD-aware fine-mapping that relaxes it. A sensitivity analysis restricted to the 354 variants around the lead signal gave the same result (H4 = 0.98). Three other POP loci that neighbour eBMD instruments behaved differently. EFEMP1 was dominated by H2 (association with eBMD only, 0.58), SMAD3 by H3 (0.99), and ESR1 showed window-dependent results (region-level H4 = 0.12; S4 Table). 18q21.1 was therefore distinctive among the loci examined.

Regional colocalisation analysis suggested overlapping association signals at the SMAD7 locus for POP and eBMD; however, fine-mapping via SuSiE resolved two distinct, moderately linked credible sets (rs4939832 for POP and rs2337106 for eBMD, r^2^ = 0.21, separated by 4.7 kb), arguing against a single shared causal variant. LD-aware fine-mapping sharpened this regional picture. We ran SuSiE independently on both traits over the same 2,473 variants, using a 1000 Genomes EUR LD matrix (n = 503). POP returned a single 95% credible set of four variants led by rs4939832 (posterior inclusion probability 0.36). Its strongest proxy rs9955731 (r^2^ = 0.95) joined the same set. eBMD returned rs2337106 as the sole member of its lead credible set (PIP 0.996). The two credible sets did not overlap: rs4939832 and rs2337106 lie 4.7 kb apart with r^2^ = 0.21, and rs4939832 had an eBMD PIP of 0.0007 despite its genome-wide significant eBMD association. Approximate conditional analysis agreed.

Conditioning rs4939832 on rs2337106 reduced its eBMD z-score from 8.07 to 3.9, whereas rs2337106 retained z = 7.0 (S1 Fig; S2 Fig; S1 Text). The contrast between the two methods is itself informative: two neighbouring variants in weak-to-moderate LD can inflate the H4 posterior probability under the single-causal-variant assumption, whereas SuSiE separates the signals. Simulation work pairing coloc with SuSiE has demonstrated more accurate inference when multiple causal variants lie in proximity [8], and fine-mapping takes primacy over regional colocalisation for biological interpretation at this locus.

Under the two-signal reading, the eBMD association observed at rs4939832 is largely driven by LD with the eBMD signal at rs2337106, and the POP association of rs2337106 is absent (conditional z = 3.1).

As a complementary view of the same locus, we performed a local genetic correlation analysis of POP and eBMD across the 1.6 Mb region using 4,583 variants [9]. The estimate was positive, consistent with partial local genetic covariation between POP and eBMD, whereas a control window containing no genome-wide significant signal returned an estimate compatible with zero. The in-house estimator is unbounded, and the result is interpreted only qualitatively, with full numerical detail provided in S1 Text.

### 2.4 Replication assessment across independent POP GWAS resources

We queried rs4939832 in two further POP resources. In the UK Biobank round-2 GWAS of genital prolapse (ICD-10 N81; n = 361,194, both sexes, linear model) [10], the same allele showed an association in the same direction. The estimate was β = 7.4 × 10^−4^ (SE 3.9 × 10^−4^; z = 1.90; two-sided P = 0.058).

This analysis used a linear model, so effect sizes are not directly comparable with FinnGen. Two nearby variants were nominally stronger (chr18:46,463,756, P = 5.9 × 10^−4^; chr18:46,463,136, P = 7.5 × 10^−4^; GRCh37 coordinates), so the UKB lead does not coincide with the FinnGen lead. We therefore describe this as a region-level, same-direction signal of nominal strength, not a replication of the variant (S3 Table).

In the largest POP GWAS to date (Iceland and UK Biobank; 15,010 cases with hospital-based diagnosis and 340,734 female controls) [3], rs4939832 was never tested or reported. It does not appear in any of the 29 supplementary tables, and none of the reported genome-wide significant loci maps to chromosome 18. The nearest tested variant, rs3764482, lies 3.3 kb away (CEU r^2^ = 0.043) and was nominally non-significant (OR = 1.028, P = 0.085). The 18q21.1 signal is therefore not yet supported outside the Finnish cohort under currently available summary-level resources. This could reflect Finnish-enriched allele effects, phenotypic heterogeneity across cohorts, or the fact that rs4939832 was not assayed in the Iceland–UK GWAS.

### 2.5 rs4939832 is not associated with female pelvic geometry

Because rs4939832 lies in SMAD7, one plausible model is mediation through pelvic skeletal geometry. We queried the variant in 21 pelvic-phenotype GWAS derived from UK Biobank DXA scans (7 phenotypes, each analysed in combined, female and male strata) [11]. No association passed the study-wise Bonferroni threshold of 2.4 × 10^−3^. All seven female strata returned null results (P = 0.13–0.63).

Nominally significant associations occurred only in the combined and male strata (P = 0.0033–0.027). The strongest was pelvic width in males (P = 0.0033), and the combined stratum contributed four nominal signals (P = 0.0075–0.027), a number compatible with chance at α = 0.05 (S2 Table). Collectively, these findings provide no supportive evidence for rs4939832 acting through female pelvic skeletal geometry; however, limited statistical power means a pelvic-geometry-mediated pathway cannot be definitively excluded.

### 2.6 No detectable cis-eQTL effect on SMAD7 in blood, immune cells or target-adjacent tissues

#### #### 2.6.1 Blood-derived and immune-cell cis-eQTL results

A simple biological model predicts that the POP risk allele alters SMAD7 expression. We queried rs4939832 against SMAD7 in the eQTL Catalogue [12] across 28 blood and immune-cell analyses spanning seven independent donor panels [13–16] (Table 2; Fig 2; S1 Table). Sample sizes ranged from 82 to 800. No analysis reached nominal significance (minimum P = 0.118, TwinsUK whole blood). The fixed-effect pooled estimate was near zero (β = −0.0036, SE 0.0129, P = 0.78), with no heterogeneity beyond expectation (Cochran Q = 16.3, df = 27). Analyses within the same donor panel share participants and are statistically non-independent. Collapsing to panels left three negative and four positive panel-level estimates (Stouffer Z = −0.47, P = 0.64). Large cohorts argue against insufficient power: GTEx v10 whole blood (n = 800) returned P = 0.999 [14], and OneK1K CD14^+^ monocytes (n = 560) returned P = 0.899 [15].

**Table 2.** cis-eQTL association of rs4939832 with SMAD7, by donor panel.

| **Donor panel** | **Analyses** | **n range** | **Panel-level β (per G)** | **Panel-level z** | **Direction** |
|---|---|---|---|---|---|
| Alasoo 2018 (macrophage, 4 conditions) | 4 | 84 | +0.0125 | +0.20 | + |
| BLUEPRINT (monocyte, neutrophil, CD4<sup>+</sup> T) | 3 | 167–196 | +0.0446 | +0.80 | + |
| GTEx whole blood (v8; v10) | 2 | 670; 800 | +0.0039 | +0.24 | + |
| Lepik 2017 (whole blood) | 1 | 471 | −0.0126 | −0.22 | − |
| OneK1K (CD14<sup>+</sup>; CD16<sup>+</sup> monocyte) | 2 | 373–560 | +0.0491 | +0.97 | + |
| Schmiedel 2018 (15 immune-cell types) | 15 | 82–91 | −0.0540 | −1.67 | − |
| TwinsUK (whole blood) | 1 | 195 | −0.1392 | −1.57 | − |
| **Summary** | **28** | **82–800** | **−0.0036 (fixed-effect)** | **−0.28 (P = 0.78)** | **4 + / 3 − (Stouffer Z = −0.47, P = 0.64)** |

Panel-level values are inverse-variance fixed-effect estimates within each panel. Panel-level directions are descriptive and must not be read as a directional signal. Minimum single-analysis P = 0.118 (TwinsUK). The full 28-row table is provided in S1 Table.

**Figure 2.**
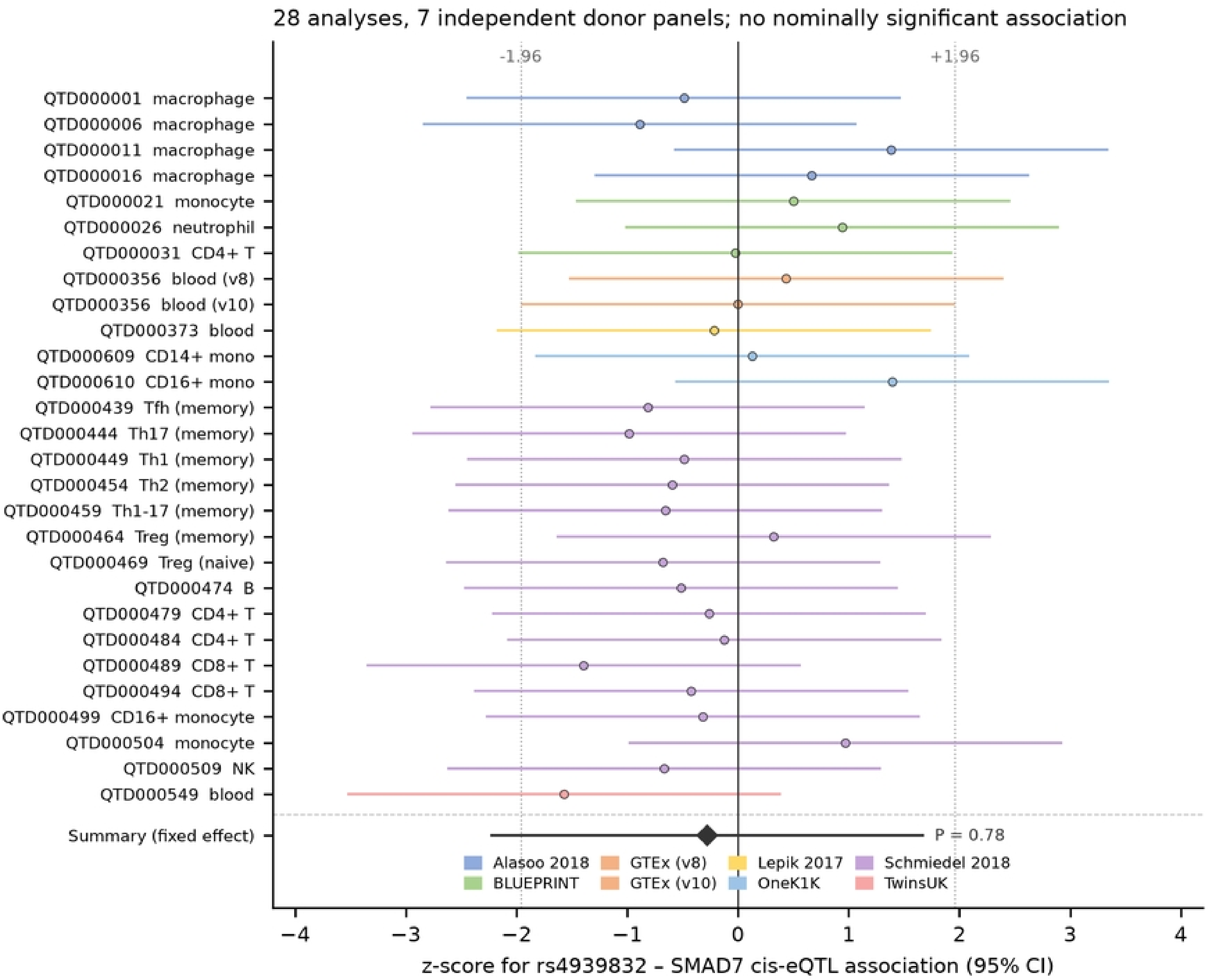
Forest plot of cis-eQTL associations between rs4939832 and SMAD7 across 28 blood-derived analyses. Summary meta-analytic effect estimate is shown at the bottom. Each row is one analysis from the eQTL Catalogue, coloured by donor panel; lines show 95% confidence intervals of the z-score, and no analysis crosses nominal significance. The seven donor panels are annotated in Table 2, and analyses within a panel share participants and are therefore not statistically independent. Corresponding cis-eQTL results from GTEx v8 vagina and uterus are provided in S6 Table.

The query pipeline was validated in two ways. Every dataset returned records for 9–11 genes at the queried position, confirming that the variant was tested rather than absent from the imputed panel. A full-window query in BLUEPRINT monocytes detected 113 SMAD7 cis-eQTL records with a region-wide best P = 0.0091 [13], confirming that the pipeline can detect real SMAD7 signals.

#### #### 2.6.2 cis-eQTL in pelvic-adjacent bulk tissues (GTEx vagina and uterus)

Two further resources addressed the remaining possibilities. SMAD7 is not measured in the deCODE SomaScan v4 plasma-proteome panel [17]. Circulating-protein mediation therefore cannot be assessed with public data. The closest available tissues are GTEx vagina and uterus. SMAD7 is highly expressed in both (median TPM 33 and 29) [14], so we tested rs4939832 directly in the GTEx v8 cis-eQTL release rather than inferring regulation from expression alone. Neither tissue yielded a significant cis-association (vagina: β = −0.124, SE 0.110, P = 0.26; uterus: β = 0.034, SE 0.114, P = 0.77; n = 141 and 129). In total, 2,638 and 2,536 distinct variants were tested, respectively. No variant in either tissue reached genome-wide significance within the SMAD7 region (regional minimum P = 1.1 × 10^−3^ and 1.5 × 10^−3^; S6 Table). The two point estimates fall on opposite sides of zero, but both confidence intervals include it, so these data constrain neither the magnitude nor the direction of any effect. We note that bulk vagina and uterus are proxies and do not represent specialised pelvic-floor connective-tissue compartments.

### 2.7 Reverse MR of POP on eBMD is not interpretable

As a negative-control sensitivity test only, we performed reverse MR (POP → eBMD) using 30 independent GWAS lead variants with strong instrument strength (F-statistic ≥ 30; median 39). Outcome summary statistics were available for 21 of the 30 leads. The IVW association reached genome-wide significance (β = 0.031, P = 1.6 × 10^−9^). However, extreme heterogeneity (Cochran Q = 1337, df = 20) indicated that the estimate was driven exclusively by two outlier SNPs (rs851996, rs4939832). The remaining 19 instruments showed null and inconsistent effect sizes. We therefore deem this reverse MR result non-interpretable for causal inference. rs851996 lies in the TNFRSF11B (osteoprotegerin) region and is one of the strongest eBMD loci, so the POP instruments are entangled with bone biology by construction. The other outlier, rs4939832, is the POP lead variant studied throughout this manuscript; an instrument associated with both traits violates the exclusion restriction by construction, which further precludes causal interpretation of this estimate.

## 3. Discussion

### 3.1 Locus-restricted overlap, not a systemic bone pathway

Our results localise the genetic overlap between POP and eBMD to individual loci. At the global level, our companion MR analysis excludes a causal effect of genetically predicted eBMD on POP larger than about 10% per SD (S5 Table). At the locus level, however, seven POP loci lie within 500 kb of an eBMD instrument. Among them, 18q21.1 shows the cleanest alignment: the same allele (G) associates with higher POP risk and higher eBMD. The locus shows regional colocalisation under the single-causal-variant assumption; fine-mapping resolves two adjacent independent causal signals with no overlapping credible sets. To our knowledge it is the only POP locus where a single variant reached genome-wide significance for both traits in released summary statistics.

This dissociation — no global causal effect despite local signal sharing — indicates that the 18q21.1 association is not mediated through systemic bone mass. Whether either causal variant acts in pelvic-floor connective tissue, or in bone with a secondary local effect, cannot be decided with current data. The high regional H4 probability output by the coloc.abf method arises from LD between two neighbouring but distinct causal variants, violating the method’s single-causal-variant assumption. Regional colocalisation here therefore serves as a screening statistic, and the fine-mapping result takes primacy for biological inference [8]. The two variants are 4.7 kb apart, so both plausibly mark one regulatory domain; assigning each to a tissue-specific mechanism requires functional data that do not yet exist.

### 3.2 SMAD7 remains a hypothesis, not a mechanism

SMAD7 is an intracellular inhibitor of TGF-β signalling, and altered TGF-β/Smad-pathway protein abundance has been reported in prolapsed vaginal wall [5]. These observations make SMAD7 a biologically plausible candidate at 18q21.1. Our eQTL analysis, however, provides no support for a simple model in which rs4939832 tunes SMAD7 expression in circulating immune cells. We emphasise the limits of that inference. Blood eQTL panels are dominated by immune cells. No dedicated pelvic floor eQTL resource exists, and the closest proxies available to us, bulk GTEx vagina and uterus, returned null. A regulatory effect confined to fibroblasts or smooth-muscle cells of the vaginal wall would be diluted in bulk tissue and therefore remains untested. These protein-level observations obtained from diseased prolapse tissue [5] cannot be directly attributed to the germline genetic variant investigated here, because they cannot separate downstream molecular changes arising from established disease from upstream genetically driven perturbations.

### 3.3 Population specificity and phenotype definition

The 18q21.1 signal reached genome-wide significance only in FinnGen. UK Biobank showed a same-direction, region-level signal at nominal strength, with a different lead variant. The Iceland–UK GWAS did not test the variant at all. Three explanations are compatible with these observations: founder-effect enrichment of the signal in Finland [6]; heterogeneity of POP phenotyping across register-based and surgical definitions [3]; and limited power of the comparison resources. We do not distinguish between them. We therefore caution against describing 18q21.1 as a general POP locus.

### 3.4 Limitations

Cis-eQTL analyses across 28 blood-derived datasets yielded no statistically significant association between rs4939832 and SMAD7 expression (meta-analytic P = 0.78); these analyses span seven independent donor panels, with sample sizes ranging from 82 to 800, and analyses within a panel share participants and are therefore statistically non-independent. Interrogating pelvic floor-adjacent tissues from GTEx v8 (vagina, n = 141; uterus, n = 129) likewise returned no significant cis-association (P = 0.26 and 0.77, respectively). These observations suggest that any cis-regulatory effect of rs4939832 on SMAD7 expression is likely small, and its directionality cannot be reliably inferred from available tissue resources.

Several further limitations apply. The local genetic correlation was estimated with an in-house implementation because the reference tool could not be compiled, the estimator is unbounded, and its confidence interval is jackknife-based [9]. The UKB POP phenotype is register-based (ICD-10 N81) and includes milder and possibly incidental diagnoses [3,10]. The eBMD and UKB POP analyses share the UK Biobank cohort, so the secondary replication is not fully independent of the exposure GWAS. No dedicated pelvic floor eQTL resource exists, and the closest proxies, GTEx vagina (n = 141) and uterus (n = 129), were interrogated here and returned null but are modestly powered and cannot exclude a small effect. Although SuSiE fine-mapping separated adjacent POP- and eBMD-associated credible sets, LD between credible-set members leaves residual uncertainty about whether shared functional regulatory elements exist in the locus (maximum cross-trait r^2^ = 0.32, far below the ≈0.8 at which proxies typically merge). Finally, our study concerns one locus, making no claims about the other POP loci that neighbour eBMD instruments. Reverse-MR results are non-interpretable, partly because rs4939832 itself acts as one of the POP genetic instruments.

### 3.5 Overlap with a related manuscript

The FinnGen R12 POP GWAS is also analysed in a manuscript on the PCOLCE2 locus submitted elsewhere by our group. That manuscript also contains the global MR and equivalence analysis summarised in S5 Table. The two manuscripts share the outcome GWAS but no analyses, no loci, and no conclusions. PCOLCE2 maps to chromosome 3, and no signal discussed here involves chromosome 3. No variant described in the two manuscripts overlaps. We will disclose this relationship at submission.

## 4. Methods

### 4.1 Data sources

We used FinnGen R12 release summary statistics for N14_FEMGENPROL (female genital prolapse; 23,074 cases, 130,160 controls; GRCh38) [6]. Case and control counts were taken from the FinnGen R12 endpoint metadata. The number of variants at P < 5 × 10^−8^ was recounted from the released file. eBMD summary statistics were from Morris et al. (GCST006979; n = 426,824; GRCh37) [4]. UK Biobank round-2 N81 summary statistics were from the Neale lab resource (n = 361,194; GRCh37) [10]. Pelvic-phenotype GWAS were from Xu et al. (GCST90448906–90448926) [11]. Summary statistics for the Iceland–UK POP GWAS were taken from its published supplementary tables [3]. eQTL summary statistics were from the eQTL Catalogue releases 7 and 8 beta [12].

### 4.2 Locus definition and harmonisation

Variants with P < 5 × 10^−8^ in the FinnGen POP GWAS were merged within ±500 kb into 30 loci. eBMD instruments were mapped to each locus window. For rs4939832, alleles were harmonised by effect allele: FinnGen reports the ALT allele as the effect allele, and the eBMD statistics were flipped to this convention. Effect alleles were cross-checked against frequencies (FinnGen 0.255; eBMD 0.242; UKB 0.2426; 1000 Genomes CEU 0.212) before any comparison. A and G are not complementary bases, so strand flips cannot convert one label into the other. Frequency agreement was therefore required rather than assumed. For eBMD, published P values were retained rather than recomputed. In the released file the reported P and the value implied by β and SE diverge for strong signals. Therefore β/SE-derived z-scores are used only for the locus figure, and published P values are used for all inference.

### 4.3 Local genetic correlation

We estimated ρ across the 1.6 Mb window around rs4939832 using summary-statistic z-scores and a 1000 Genomes EUR Phase 3 LD matrix (489 haploid-equivalent samples; hg19) [9]. The estimator follows the LAVA formulation of local genetic covariance [9]. Standard errors were obtained by delete-one-block jackknife over 20 blocks. A control window (chr18:30.0–31.6 Mb) with no genome-wide significant signal in either trait served as a calibration check. Full numerical detail is in S1 Text.

### 4.4 eQTL queries

For each eQTL Catalogue dataset we queried an 11 bp window around rs4939832 (GRCh38 chr18:48,939,295) from remote tabix indices [12]. Queries that failed at this width were retried with a 30 kb window and positional filtering. The effect allele was ALT (G) in all datasets. We extracted β, SE, P and MAF for SMAD7 (ENSG00000101665) at the queried position. We pooled estimates with inverse-variance weighting and combined z-scores with Stouffer’s method, unweighted and √n-weighted. Heterogeneity was assessed with Cochran’s Q. Donor-panel membership was annotated for every analysis and used for the panel-level summary.

### 4.5 Fine-mapping and credible-set colocalisation

SuSiE [18] was run independently for POP and eBMD over 2,473 variants at the locus, using summary-statistic z-scores and L = 10 latent signals with no residual-variance estimation. The LD correlation matrix came from 1000 Genomes Phase 3 EUR (n = 503; hg19 positions). Positions were lifted from GRCh38 to GRCh37 with the UCSC hg38ToHg19 chain and matched to the 1000 Genomes EUR reference panel by position; alleles were harmonised. Variants that could not be matched after liftOver or were monomorphic in the reference panel were dropped (18 of 2,491 input variants), leaving 2,473 variants for fine-mapping. Credible sets were called at 95% coverage. Conditional z-scores were approximated as z(A | B) = (z_A − r z_B)/√(1 − r^2^), where r is the LD correlation between the two variants. PIPs, credible sets and pairwise LD values are in S1 Text (S1 and S2 Figs).

### 4.6 Reverse MR

Thirty independent POP lead variants were defined by merging genome-wide significant variants within 500 kb and keeping the most significant variant per block. Instruments were matched to the Morris eBMD resource by rsID with allele harmonisation (21 available). Per-variant Wald ratios, fixed-effect IVW, Cochran Q and leave-one-out estimates were computed with first-order weights.

### 4.7 Software

Analyses used Python 3.13 with numpy, pandas, pyarrow and matplotlib. The local-correlation and jackknife code will be deposited in the repository described in Data availability.

## Data availability

All GWAS and eQTL summary statistics analysed in this work are publicly available third-party datasets: FinnGen R12 (https://www.finngen.fi/en/access_results), GWAS Catalog accessions GCST006979 (Morris et al. eBMD), GCST90448906–90448926 (Xu et al. pelvic phenotypes) and GCST010174 (Olafsdottir et al. POP), the Neale Lab UK Biobank round-2 resource (http://www.nealelab.is/uk-biobank), the eQTL Catalogue (https://www.ebi.ac.uk/eqtl) and GTEx v8 and v10 (https://gtexportal.org). We cannot redistribute these third-party summary statistics due to data-use constraints. No individual-level data were used. Custom analysis code generated in this study will be deposited in a public GitHub repository upon manuscript acceptance.

## Ethics statement

This study used only published summary statistics. The source cohorts were approved by their respective ethics bodies, and participants provided informed consent as described in the source publications [3,4,6,10,11].

## Funding

This research received no external funding.

## Competing interests

The authors declare no competing interests.

## Author contributions

**CRediT taxonomy:** conceptualization, S.H. and J.T.; methodology, J.T. and S.H.; software, J.T. and S.H.; validation, J.T., H.L. and S.H.; formal analysis, H.L., S.H. and J.T.; investigation, J.T. and H.L.; data curation, J.T. and H.L.; writing—original draft preparation, J.T. and S.H.; writing—review and editing, J.T.; visualization, S.H. and J.T.; supervision, S.H.; project administration, J.T. and S.H. J.T., Jinhua Tian; H.L., Hongming Li; S.H., Songming He.

## Acknowledgements

We acknowledge the participants and investigators of the FinnGen study and UK Biobank.

## Figure legends

**S1 Fig. Credible-set posterior inclusion probabilities for POP and eBMD at 18q21.1**.

Each point is one of 2,473 variants. Axes show SuSiE PIPs from the trait-specific analyses. POP credible-set members (rs4939832, rs9955731, rs1873190, rs1873191) are filled circles; the eBMD lead rs2337106 is a diamond. The two credible sets do not overlap.

**S2 Fig. LD r**^**2**^ **between the POP credible set and the eBMD lead variant**.

Bars show pairwise r^2^ from 1000 Genomes EUR (n = 503), computed between rs2337106 and each member of the POP credible set, and within the POP credible set (rs4939832–rs9955731 shown for reference). The lead-to-lead r^2^ is 0.21. The largest cross-set r^2^ is 0.32 (rs1873190 and rs1873191, in perfect LD with each other, versus rs2337106), still far below the r^2^ ≈ 0.8 at which proxies typically merge into one credible set; the pairwise values underlying this figure are given in S1 Text (S1.11), and the full 4 × 281 cross-set LD matrix (4 POP credible-set members against all 281 eBMD credible-set members at this locus) is provided by the analysis outputs described in S1 Text (S1.14).

## Supporting information

- **S1 Text**. Local genetic correlation at 18q21.1 (estimator, windows, three-tier implementation verification, calibration and the direct allelic evidence) and the colocalisation method comparison (single-causal-variant assumption of coloc.abf, SuSiE credible sets, conditional z-scores and pairwise LD values).
- **S1 Table**. cis-eQTL association of rs4939832 with SMAD7 across 28 blood-derived analyses, with donor-panel annotation.
- **S2 Table**. rs4939832 in 21 pelvic-phenotype GWAS (Xu et al. 2025), by sex stratum.
- **S3 Table**. Region-level evidence in independent POP resources (UKB N81 neighbouring variants; Iceland–UK GWAS region coverage).
- **S4 Table**. Colocalisation analysis (coloc.abf) at four POP loci that neighbour eBMD instruments: posterior probabilities for H0–H4, region-level and lead-signal-level analyses.
- **S5 Table**. Global two-sample MR of eBMD on POP: main and sensitivity estimates with TOST equivalence bounds.
- **S6 Table**. rs4939832–SMAD7 cis-association in pelvic floor-adjacent tissue (GTEx v8 vagina and uterus): lead-variant estimates, variants tested, and regional minimum P.
- **S1 Fig**. Credible-set PIP scatter for POP versus eBMD at 18q21.1.
- **S2 Fig**. LD r^2^ between the POP credible set and the eBMD lead rs2337106 (lead-to-lead r^2^ = 0.21; maximum cross-set r^2^ = 0.32).

## Notes

### Competing Interest Statement

The authors have declared no competing interest.

### Clinical Trial

N/A

### Author Declarations

This study only analyzed publicly available, de-identified GWAS summary statistics and gene expression datasets. No new human participants or tissue samples were collected for this analysis. All original studies generating these public datasets have obtained appropriate ethical approval and informed consent from participants.

